# Urine tenofovir and dried blood spot tenofovir diphosphate concentrations and viraemia in people taking efavirenz and dolutegravir based antiretroviral therapy

**DOI:** 10.1101/2023.09.27.23296217

**Authors:** Jienchi Dorward, Katya Govender, Pravikrishnen Moodley, Richard Lessells, Natasha Samsunder, Yukteshwar Sookrajh, Thomas R. Fanshawe, Philip J. Turner, Christopher C. Butler, Paul K. Drain, Gail N. Hayward, Nigel Garrett

**Affiliations:** Nuffield Department of Primary Care Health Sciences, University of Oxford, Oxford, UK; Centre for the AIDS Programme of Research in South Africa (CAPRISA), University of KwaZulu–Natal, Durban, South Africa; Africa Health Research Institute, Durban, South Africa; KwaZulu-Natal Research and Innovation Sequencing Platform (KRISP), University of KwaZulu-Natal, Durban, South Africa; Department of Virology, University of KwaZulu-Natal and National Health Laboratory Service, Inkosi Albert Luthuli Central Hospital, KwaZulu-Natal, South Africa; eThekwini Municipality Health Unit, Durban, South Africa; Department of Global Health, Schools of Medicine and Public Health, University of Washington, Seattle, USA; Department of Medicine, School of Medicine, University of Washington, Seattle, USA; Department of Epidemiology, School of Public Health, University of Washington, Seattle, USA; Discipline of Public Health Medicine, School of Nursing and Public Health, University of KwaZulu-Natal, Durban, South Africa

**Keywords:** HIV, adherence, tenofovir, tenofovir diphosphate, viraemia, antiretroviral therapy, dolutegravir

## Abstract

**Objective:** We aimed to determine whether urine tenofovir (TFV) and dried blood spot (DBS) tenofovir diphosphate (TFV-DP) concentrations are associated with concurrent HIV viraemia.

**Design:** Cross-sectional study among people with HIV (PWH) receiving tenofovir disoproxil fumarate (TDF)-based antiretroviral therapy (ART).

**Methods:** We used dual tandem liquid chromatography and mass spectrometry to measure urine TFV and DBS TFV-DP concentrations, and evaluated their associations with concurrent viraemia ≥ 1000 copies/mL using logistic regression models. In exploratory analyses, we used receiver operating curves to estimate optimal urine TFV and DBS TFV-DP thresholds to predict concurrent viraemia.

**Results:** Among 124 participants, 68 (54.8%) were women, median age was 39 years (interquartile range [IQR] 34-45) and 74 (59.7%) were receiving efavirenz versus 50 (40.3%) receiving dolutegravir. Higher concentrations of urine TFV (1000 ng/mL increase, odds ratio [OR] 0.97 95%CI 0.94-0.99, p=0.005) and DBS TFV-DP (100 fmol/punch increase, OR 0.76, 95%CI 0.67-0.86, p<0.001) were associated with lower odds of viraemia. There was evidence that these associations were stronger among people receiving dolutegravir than among people receiving efavirenz (urine TFV p=0.072, DBS TFV-DP p=0.003). Nagelkerke Pseudo-R^2^ for the DBS TFV-DP models was higher than for the urine TFV models, demonstrating a stronger relationship between DBS TFV-DP and viraemia. Among people receiving dolutegravir, a DBS TFV-DP concentration of 483 fmol/punch had 88% sensitivity and 85% specificity to predict concurrent viraemia ≥ 1000 copies/ml.

**Conclusions:** Among PWH receiving TDF-based ART, urine TFV concentrations, and in particular DBS TFV-DP concentrations, were strongly associated with concurrent viraemia, especially among people receiving dolutegravir.

## INTRODUCTION

There is increasing interest in accurately monitoring antiretroviral therapy (ART) adherence for people with HIV (PWH). Tenofovir disoproxil fumarate (TDF) is included in fixed-dose combinations alongside emtricitabine and efavirenz, or lamivudine and dolutegravir, which are used by over 95% of people receiving ART in low- and middle-income countries[1, 2]. Therefore, objective tenofovir measurements could identify poor adherence. TDF is converted to tenofovir (TFV), which is metabolized intracellularly to tenofovir diphosphate (TFV-DP). TFV is excreted in urine and correlates with short term adherence as it has a 12-15 hour terminal half-life[3], while TFV-DP accumulates in red blood cells and correlates with medium term adherence as it has a longer half-life of 17 days[4]. Studies have shown that qualitative urine TFV[5-8] and quantitative dried blood spot (DBS) TFV-DP levels[9-11] are associated with viral suppression in PWH receiving ART, but none have compared the two measures, or determined thresholds that best predict viral suppression. Furthermore, dolutegravir has a higher genetic barrier to resistance than efavirenz, meaning that measures of adherence should be more closely associated with viral suppression, unlike efavirenz where resistance can cause viraemia despite good adherence.

Therefore, we aimed to compare the association between urine TFV, and DBS TFV-DP concentrations, with viraemia among PWH receiving dolutegravir and efavirenz-based ART. In post-hoc analyses, we also aimed to estimate optimal urine TFV and DBS TFV-DP thresholds to detect viraemia, and to assess associations between TFV levels and both HIV drug resistance (HIVDR), and self-reported adherence.

## METHODS

We conducted a cross-sectional analysis at enrolment into a randomised study of point-of-care HIV viral load (VL) testing (POwER)[12]. We included consecutively enrolled POwER participants receiving TDF as part of dolutegravir or efavirenz-based first-line ART. Eligible PWH had a pre-enrolment VL >1000 copies/mL in the past 6 weeks, without having received enhanced adherence counselling. At enrolment, participants self-reported adherence, and had urine, DBS and plasma samples taken and stored at -80°C, for retrospective testing.

We quantitated urine TFV and DBS TFV-DP concentrations using liquid chromatography and dual tandem mass spectrometry (LC-MS/MS). We tested VL using the cobas 6800 platform (Roche, Basel, Switzerland), and attempted drug resistance testing for all samples with VL ≥ 1000 copies/mL (see supplement).

We used logistic regression models to assess the relationship between the exposure of either urine TFV concentrations, or DBS TFV-DP concentrations, and the outcome of viraemia. To determine whether associations differed by ART regimen, we included a variable for ART regimen (dolutegravir versus efavirenz) in the model, with an interaction term between ART regimen and urine TFV, or DBS TFV-DP concentrations. We fitted separate models for the outcomes of viraemia ≥1000 copies/mL, and ≥50 copies/mL, as these thresholds are used in World Health Organization guidelines [1]. We compared the Nagelkerke pseudo-R^2^ of the urine TFV and DBS TFV-DP models to determine which measure was more strongly associated with viraemia. In exploratory, post-hoc analyses, we used receiver operating curves (ROCs) to estimate urine TFV and DBS TFV-DP thresholds that maximise sensitivity and specificity to predict concurrent viraemia. Lastly, we described urine TFV and DBS TFV-DP levels among people with and without HIVDR, and compared self-reported short-term and longer-term adherence with urine TFV and DBS TFV-DP levels using logistic regression and linear regression models respectively. Sample size was determined by the number of participants enrolled into POwER and receiving TDF.

We analysed data using R 4.2.0 (R Foundation for Statistical Computing, Vienna, Austria). The University of KwaZulu-Natal Biomedical Research Ethics Committee (BREC 00000836/2019) and the University of Oxford Tropical Research Ethics Committee (OxTREC 66-19) approved the study.

## RESULTS

Between August 2020-March 2022, we enrolled 124 PWH. 68 (54.8%) were women, the median age was 39 years (interquartile range [IQR] 34-45) and 74 (59.7%) were receiving efavirenz versus 50 (40.3%) receiving dolutegravir (Table S1). 23.4% self-reported missing a dose in the past 4 days, and 62.9% reported last missing a dose over four weeks before enrolment. Median time since the pre-enrolment viraemic VL was 15 days (IQR 13-21). In December 2020 we discovered that 45 participants had pre-enrolment VLs measured on a faulty analyser, with potentially false viraemic pre-enrolment results. This did not affect the enrolment viral loads used in this analysis, but meant that not all participants had recent viraemia, and so there were a higher number of participants with viral suppression at enrolment than anticipated. Therefore, enrolment, VLs were ≥ 1000 copies/mL in 44/124 participants (35.5%), 50-999 copies/mL in 23/124 (18.5%), and suppressed <50 copies/mL in 57/124 (46.0%). Among the 43 with successful HIVDR testing 24/43 (55.8%) had mutations conferring resistance to their current regimen. Among those receiving efavirenz, 23/27 (85.2%) had resistance to their current regimen, versus 1/16 (6.3%) of those receiving dolutegravir (one person with M184V mutation alone). Median urine TFV concentration was 20000 ng/mL (IQR 7280-33625), and median TFV-DP concentration was 734 fmol/punch (IQR 471-1015).

Higher concentrations of urine TFV (1000 ng/mL increase, odds ratio [OR] 0.97 95%CI 0.94-0.99, p=0.005) and DBS TFV-DP (100 fmol/punch increase, OR 0.76, 95%CI 0.67-0.86, p<0.001) were associated with lower odds of viraemia ≥ 1000 copies/mL, with similar results at ≥ 50 copies/mL (Table 1A). There was some evidence that the association between urine TFV and viraemia at 1000 copies/mL (LRT for interaction p=0.072), and between DBS TFV-DP and viraemia at both 1000 (p=0.003) and 50 (p=0.068) copies/mL, was stronger among people receiving dolutegravir than among people receiving efavirenz. There was no evidence of a difference by ART regimen in the association between urine TFV and viraemia at 50 copies/mL (p=0.797, Table 1A). Overall, at both 1000 copies/mL and 50 copies/mL thresholds, the Nagelkerke Pseudo-R^2^ for the DBS TFV-DP models was higher than for the urine TFV models, meaning there was a stronger relationship between DBS TFV-DP and viraemia (Table 1A).

**Table 1:** A) Logistic regression models of the association between urine tenofovir concentrations, and dried blood spot tenofovir diphosphate concentrations, and viraemia B) Diagnostic accuracy thresholds and areas under the curve for urine tenofovir and tenofovir diphosphate concentrations to detect viraemia

|  | Overall |  |  | Dolutegravir |  | Efavirenz |  |
| --- | --- | --- | --- | --- | --- | --- | --- |
| A) Logistic regression models |  |  |  |  |  |  |  |
|  | Odds ratio<br>(95% CI) | P | Nagelkerke<br>Pseudo-R<sup>2</sup> | Odds ratio<br>(95% CI) | P | Odds ratio<br>(95% CI) | P |
| Viraemia ≥1000 copies/mL |  |  |  |  |  |  |  |
| Urine TFV<br>concentration<br>(increase of<br>1000 ng/mL) | 0.97 (0.94-0.99) | 0.005 | 0.107 | 0.94 (0.89-0.98)* | 0.007 | 0.98 (0.95-1.01)* | 0.250 |
| DBS TFV-DP<br>concentration<br>(increase of<br>100<br>fmol/punch) | 0.76 (0.67-0.86) | <0.001 | 0.274 | 0.53 (0.35-0.71)† | <0.001 | 0.85 (0.74-0.95)† | 0.009 |
| Viraemia ≥50 copies/mL |  |  |  |  |  |  |  |
| Urine TFV<br>concentration<br>(increase of<br>1000 ng/mL) | 0.98 (0.96-1.00) | 0.041 | 0.097 | 0.98 (0.95-1.00)‡ | 0.099 | 0.98 (0.96-1.01)‡ | 0.225 |
| DBS TFV-DP<br>concentration<br>(increase of<br>100<br>fmol/punch) | 0.86 (0.78-0.93) | <0.001 | 0.188 | 0.74 (0.58-0.89)§ | 0.010 | 0.90 (0.81-0.99)§ | 0.040 |
| B) Thresholds to detect viraemia and associated areas under the curve (AUCs) |  |  |  |  |  |  |  |
|  | Threshold<br>(sens, spec<br>[%]) | AUC [%] (95% CI) |  | Threshold<br>(sens, spec) | AUC [%]<br>(95% CI) | Threshold<br>(sens, spec<br>[%]) | AUC [%]<br>(95% CI) |
| Viraemia ≥1000 copies/mL |  |  |  |  |  |  |  |
| Urine TFV<br>concentration<br>(ng/mL) | 8340 (85.0,<br>54.5) | 69.3 (58.6-80.0) |  | 3495 (76.5,<br>93.9) | 83.2 (68.3-98.1) | 20100 (59.3,<br>57.4) | 59.2 (44.6-73.8) |
| DBS TFV-DP<br>concentration<br>(fmol/punch) | 639 (68.2,<br>76.3) | 77.9 (68.9-87.0) |  | 483 (88.2,<br>84.8) | 90.1 (80.2-100) | 816 (74.1,<br>70.2) | 74.7 (62.5-87.0) |
| Viraemia ≥50 copies/mL |  |  |  |  |  |  |  |
| Urine TFV<br>concentration<br>(ng/mL) | 18350 (56.7,<br>63.2) | 62.4 (52.4-72.3) |  | 16000 (54.5,<br>70.6) | 66.8 (51.9-81.8) | 20100 (58.8,<br>60.0) | 59.5 (46.0-73.0) |
| DBS TFV-DP<br>concentration<br>(fmol/punch) | 795 (74.6,<br>68.4) | 76.4 (68.0-84.8) |  | 686 (81.8,<br>76.5) | 84.4 (73.1-95.7) | 816 (64.7,<br>70.0) | 68.3 (55.8-80.8) |

\* LRT for interaction p = 0.072, †LRT for interaction p = 0.003, ‡LRT for interaction p = 0.797, §LRT for interaction p = 0.068, † calculated as the point which minimises $(1 - \text{Sens})^2 + (1 - \text{Spec})^2$

Overall, AUCs for concurrent viraemia at 1000 copies/mL were modest (urine TFV 69.3%, DBS TFV-DP 77.9%, Table 1B). However, AUCs were higher among people receiving dolutegravir compared to efavirenz, using both urine TFV (0.83, 95%CI 0.68-0.98 versus 0.59, 95% CI 0.45-0.74, p=0.026 Figure 1A) and DBS TFV-DP (0.90, 95%CI 0.80-1.00 versus 0.75, 95%CI 0.62-0.87, p=0.059, Figure 1B). From the ROC curves among people receiving dolutegravir, a urine TFV concentration threshold of 3495 ng/mL would have 77% sensitivity and 94% specificity to predict concurrent viraemia ≥ 1000 copies/ml (Table S2). A DBS TFV-DP concentration of 483 fmol/punch would have 88% sensitivity and 85% specificity to predict concurrent viraemia ≥ 1000 copies/ml. When using a threshold of ≥ 50 copies/mL, AUCs were similar to >1000 copies/mL, except with urine TFV among people receiving dolutegravir, which performed less well (Table 1A, Figure S1), compared to a threshold of 1000 copies/mL.

**Figure 1:**
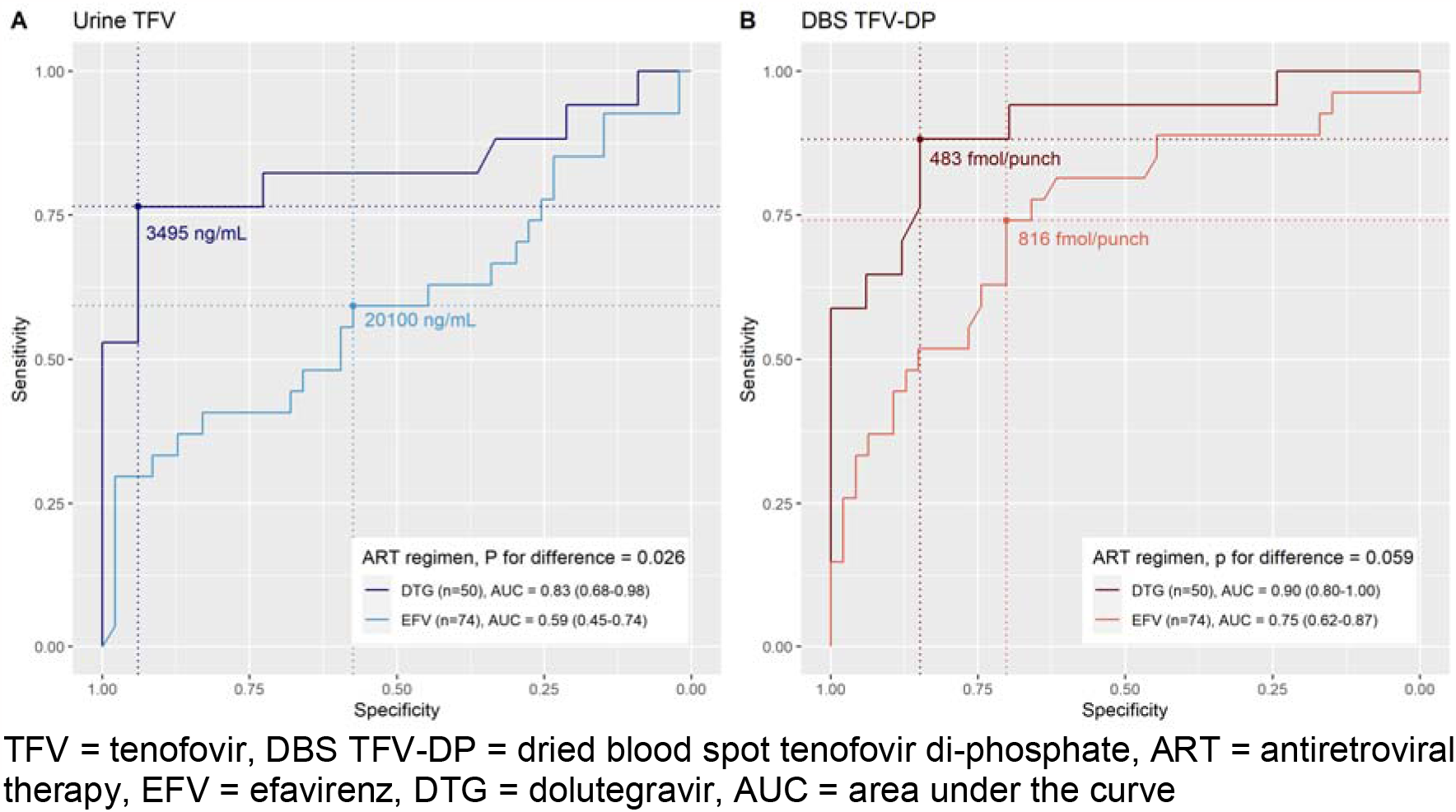
Receive operating characteristic curves of urine tenofovir and dried blood spot tenofovir diphosphate concentrations to predict viraemia ≥ 1000 copies/mL, by ART regimen.

Among 43 people with viraemia >500 copies/mL and successful HIVDR testing, median urine TFV (17300 ng/mL, IQR 1120-29350 versus 343 ng/mL, 0-20950) and DBS TFV-DP levels (646 fmol/punch, IQR 388-820 versus 103 fmol/punch, 10-374) were higher among people with HIVDR compared to those without, but numbers were too small for formal comparisons or meaningful breakdown by ART regimen (Table S3).

Self-reported missed doses in the past four days and more recently self-reported missed doses were both associated with lower urine TFV and DBS TFV-diphosphate concentrations (Table S4 and Figure S2).

## DISCUSSION

In this cross-sectional study we found that urine TFV and DBS TFV-DP concentrations were negatively associated with concurrent viraemia, and the association was generally stronger with dolutegravir compared to efavirenz. Furthermore, DBS TFV-DP had a better association with viraemia compared to urine TFV.

Our findings are similar to studies which have shown that DBS TFV-DP concentrations are associated with viraemia in PWH receiving TDF-based ART[9-11]. A study among 532 PWH in the United States found that higher DBS TFV-DP was associated with VL <20 copies/ml. 36% of participants were receiving integrase inhibitors, and 27% were receiving non-nucleoside reverse transcriptase inhibitors. Among people with VL <20 copies/mL, the mean TFV-DP concentration was 1728 (1608-1857) fmol/punch compared to 1469 (1283-1681) fmol/punch at 20-200 copies/mL, and 633 (542-739) fmol/punch at >200 copies/mL, but unlike our study results were not presented by ART class[9]. A South African cross-sectional study among 137 people taking efavirenz used ROC curves to demonstrate that DBS TFV-DP was more strongly associated than plasma TFV with viral suppression <50 copies/mL[10]. Lastly, among 250 virally suppressed PWH receiving efavirenz in South Africa, baseline TFV-DP <400 fmol/punch was associated with increased odds of developing viraemia ≥ 400 copies/mL after 1 month[11]. Regarding urine TFV levels, several studies demonstrate qualitative point-of-care urine TFV levels are associated with concurrent viraemia[5-8], but no studies have assessed the relationship between quantitative TFV concentrations and viraemia.

While both urine TFV and DBS TFV-DP concentrations were negatively associated with viraemia ≥ 1000 copies/mL, the association was weaker with efavirenz, likely because the high prevalence of HIVDR means viraemia persists in the presence of measurable adherence. With dolutegravir, HIVDR was rare, meaning the relationship between TFV measures and viraemia was stronger. In exploratory, post-hoc analyses using ROC curves, we similarly found that the potential for urine TFV and DBS TFV-DP to predict concurrent viraemia was poor to modest with efavirenz, but more acceptable with dolutegravir. Using DBS TFV-DP, a threshold of around 480 fmol/punch would have >80% sensitivity and specificity to predict concurrent viraemia ≥ 1000 copies/mL. Comparing pseudo-R^2^ values, models indicated superiority of fit for TFV-DP over urine TFV, suggesting that TFV-DP generally performed better. This is likely because quantitative DBS TFV-DP reflects longer-term adherence, which is required to achieve viral suppression, and which is not captured by more transient, shorter-term urine TFV concentrations.

Strengths of our study include the comprehensive assessment of adherence using viral load, HIVDR, self-reported adherence, urine TFV and DBS TFV-DP. To our knowledge, this is the first study to directly compare the predictive value of quantitative urine TFV concentrations against DBS TFV-DP in PWH. We used different viraemia thresholds which reflect WHO guidelines[1]. Our study is limited by the small sample size, with participants enrolled in a clinical trial for people with recent viraemia, meaning results may not be generalizable to other populations. While we estimate urine TFV and DBS TFV-DP thresholds to optimize sensitivity and specificity to detect viraemia at both ≥ 50 and ≥ 1000 copies/mL, these are exploratory analyses with imprecise estimates due to our small sample size. These thresholds should be evaluated in other studies, and should take into account the clinical use scenario, and whether sensitivity or specificity should be maximized[13]. Therefore, we provide ranges of thresholds and associated sensitivities and specificities in Table S2.

Our study suggests that urine TFV or DBS TFV-DP measures of adherence may be increasingly useful given the global dolutegravir rollout. Qualitative point-of-care urine TFV assays have been validated[5-8], and their clinical effectiveness is being investigated in clinical trials[14]. While TFV-DP performed better in our study, it currently requires expensive LC-MS/MS. Development of point-of-care TFV-DP assays should be prioritized[15], alongside studies to establish if there is a reliable TFV-DP threshold for predicting concurrent viraemia among people receiving dolutegravir.

## Data Availability

Bona fide researchers will be able to request access to anonymised trial data by contacting the corresponding author.

## LIST OF ABBREVIATIONS

ART –: antiretroviral therapy
AUC –: area under the curve
DBS –: dried blood spot
HIVDR –: HIV drug resistance
LC-MS/MS–: liquid chromatography and dual tandem mass spectrometry
PWH –: people with HIV
ROC –: receiver operating characteristic
TDF –: tenofovir disoproxil fumarate
TFV –: tenofovir
TFV-DP –: tenofovir diphosphate
VL –: viral load

## DECLARATIONS

### Competing interests

The authors have no competing interests to declare.

## Funding

This work is supported by grants from the Wellcome Trust PhD Programme for Primary Care Clinicians (216421/Z/19/Z) and the University of Oxford’s Research England QR Global Challenges Research Fund (0007365). HIV drug resistance testing and drug concentration testing was funded by the National Institute for Health and Care Research (NIHR) Community Healthcare MedTech and In Vitro Diagnostics Co-operative at Oxford Health NHS Foundation Trust (MIC-2016-018); GH, CCB & PJT also receive funding from this award. JD, Academic Clinical Lecturer (CL-2022-13-005), is funded by the UK National Institute of Health and Social Care Research (NIHR). The views expressed are those of the author(s) and not necessarily those of the NHS, the NIHR or the Department of Health and Social Care. For the purpose of open access, the author has applied a CC BY public copyright licence to any Author Accepted Manuscript version arising from this submission.

## Author contributions

JD, PKD and NG conceived the study. KG, PM, RL and NS were responsible for laboratory testing. RL, YS, PT, CB and GH contributed to study design. JD analysed the data with guidance from TF. JD wrote the first draft of the manuscript. All authors critically reviewed and edited the manuscript and consented to final publication.

## Acknowledgements

The authors would like to thank all participants in the study and acknowledge the work and support of staff at the Prince Cyril Zulu Clinic, Mafakathini Clinic, eThekwini Municipality, CAPRISA, AHRI Pharmacology Core, KRISP and the National Health Laboratory Services at Inkosi Albert Luthuli Hospital. Thank you to Ms Lara Lewis for statistical input.

## Notes

### Competing Interest Statement

The authors have declared no competing interest.

### Author Declarations

The University of KwaZulu-Natal Biomedical Research Ethics Committee (BREC 00000836/2019) and the University of Oxford Tropical Research Ethics Committee (OxTREC 66-19) approved the study.

