## supplement for "Urine tenofovir and dried blood spot tenofovir diphosphate concentrations and viraemia in people taking efavirenz and dolutegravir based antiretroviral therapy"

**SUPPLEMENTARY FILE**

### 1. Liquid chromatography and dual tandem mass spectrometry (LC-MS/MS) sample analysis protocol

We conducted LC-MS/MS at the Africa Health Research Institute in Durban, South Africa. A quantitative LC-MS/MS method was developed for the determination of tenofovir (TFV) in urine samples and TFV and tenofovir-diphosphate (TFV-DP) concentrations in dry blood spot (DBS) samples. The LC-MS/MS method was accurate, robust and quantitative over the concentration ranges; 0.5 – 80 µg/mL for TFV in urine and 100 – 8000 pg/mL for TFV and TFV-DP in DBS samples.

The urine and DBS samples were processed using a protein precipitation method. A 70% methanol:water (v/v) solution, which contained the deuterated internal standards; d6-TFV and d5-TFV-DP was used for drug analyte extraction. The calibration standards and quality control solutions (containing TFV and TFV-DP) were prepared using the extraction solution.

The LC-MS/MS analysis was performed using an Agilent high pressure liquid chromatography (HPLC) system coupled to an AB Sciex 5500, triple quadrupole mass spectrometer equipped with an electrospray ionization (ESI) TurboIonSpray source. Analyst software, version 1.6.2 was used for data acquisition and quantitative data analysis.

Tenofovir and TFV-DP was quantitated using ion pair-hydrophilic interaction chromatography coupled to tandem mass spectrometry (IP–HILIC–MS/MS). The chromatographic separation was performed at a flow rate of 0.2 mL/min on a Luna Amino (NH2) column (Phenomenex, Torrance, CA) 100 mm × 2.0 mm, packed with 3.0 µm particles. Mobile phase A consisted of 100 mM hexafluoro-2-propanol (HFIP) and 0.5% diethylamine (DEA) (v/v) in water, and mobile phase B consisted of 0.1 M HFIP and 0.5% DEA (v/v) in acetonitrile. A sample volume of 5.0 µL was injected onto the HPLC column and the analytes were separated using a gradient elution. The autosampler syringe and the injection valve were washed with a water:acetonitrile (30:70, v/v) solution, post sample injection, to reduce carryover. The system was operated in negative-ion multiple reaction monitoring (MRM) mode set to detect precursor [M+H]^+^→ product ion transitions for TFV1 (*m/z* 285.8 → *m/z* 133.9), TFV2 (*m/z* 285.8→ *m/z* 151.0), TFV-DP1 (*m/z* 445.8 → *m/z* 158.9), TFV-DP2(*m/z* 445.8→ *m/z* 176.7) and the internal standard; d6-TFV (*m/z* 292.0→ *m/z* 133.8) and d5-TFV-DP (*m/z* 450.8→ *m/z* 158.9). The optimized ESI source dependent parameters were set as follows; ion spray voltage (ISV): 5500V, temperature (TEM): 350°C, gas 1 (N_2_) and gas 2 (N_2_): 40 psi.

### 2. Details of HIV viral load testing

We tested viral laod with the cobas® HIV-1 assay using the cobas 6800 platform (06998836190 ; Roche, Basel, Switzerland) at the Inkosi Albert Luthuli Hospital in Durban, South Africa.

### 3. Details of HIV drug resistance testing

HIV drug resistance testing was conducted at the KwaZulu-Natal Research Innovation and Sequencing Platform in Durban, South Africa. We attempted sequencing of HIV-1 *pol* (protease [PR], reverse transcriptase [RT] and integrase [IN]) for all samples with VL ≥1000 copies/mL. Following RNA extraction, we amplified PR, RT and IN genes using the amplification module of the Applied Biosystems HIV-1 Genotyping Kit with Integrase (Thermo Fisher Scientific, Waltham, MA); and sequenced on the Illumina MiSeq platform (Illumina, San Diego, CA). We identified drug resistance mutations (DRMs) at >20% frequency using Stanford HIVdb (version 9.1).

### 4. Figure S1 Receive operating characteristic curves of urine tenofovir and dried blood spot tenofovir diphosphate concentrations to predict viraemia ≥50 copies/mL, by ART regimen

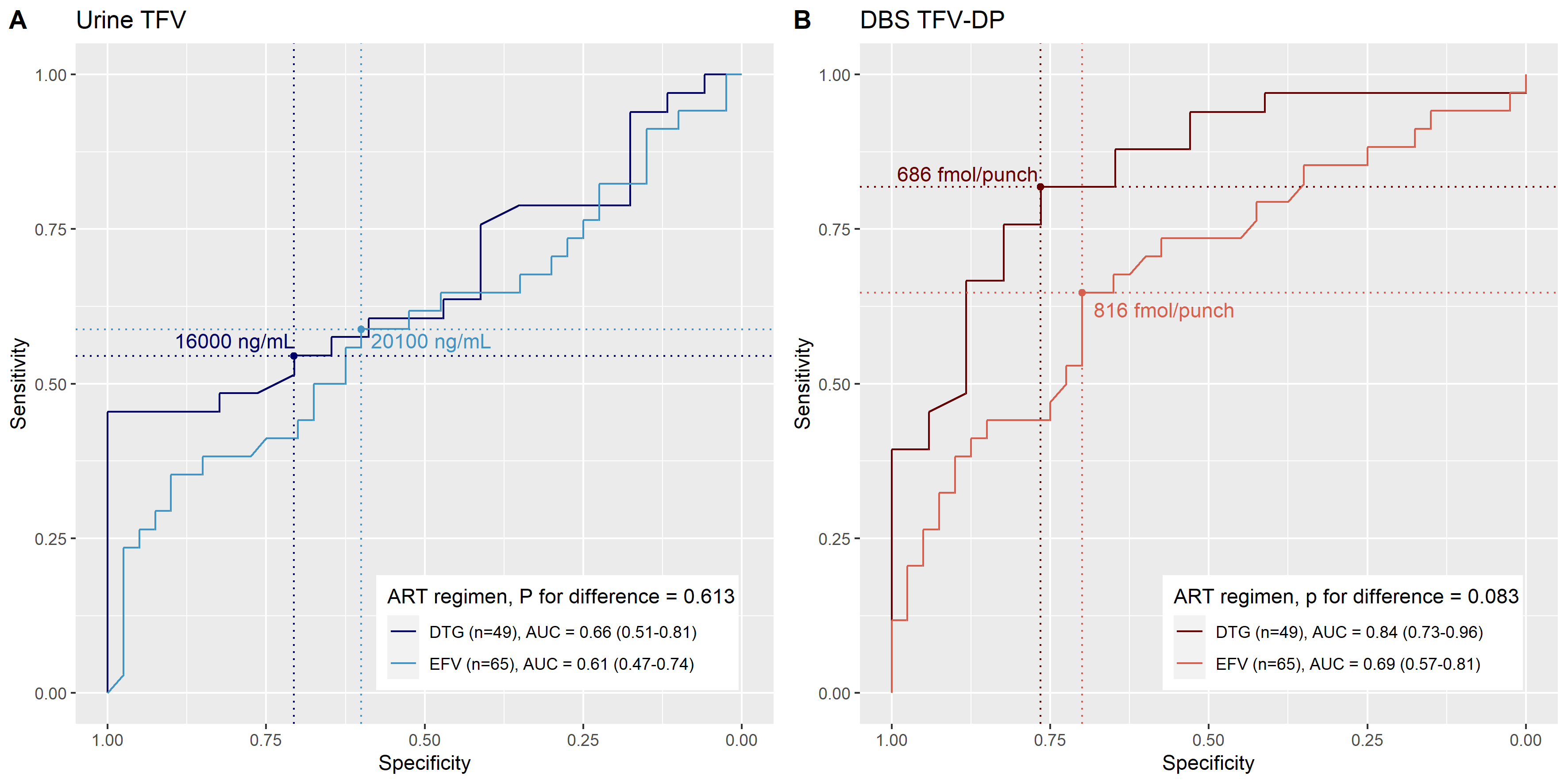

TFV = tenofovir, DBS TFV-DP = dried blood spot tenofovir di-phosphate, ART = antiretroviral therapy, EFV = efavirenz, DTG = dolutegravir, AUC = area under the curve

### 5. Figure S2 Linear regression models of the association between self-reported adherence measures and urine tenofovir and dried blood spot tenofovir diphosphate concentrations

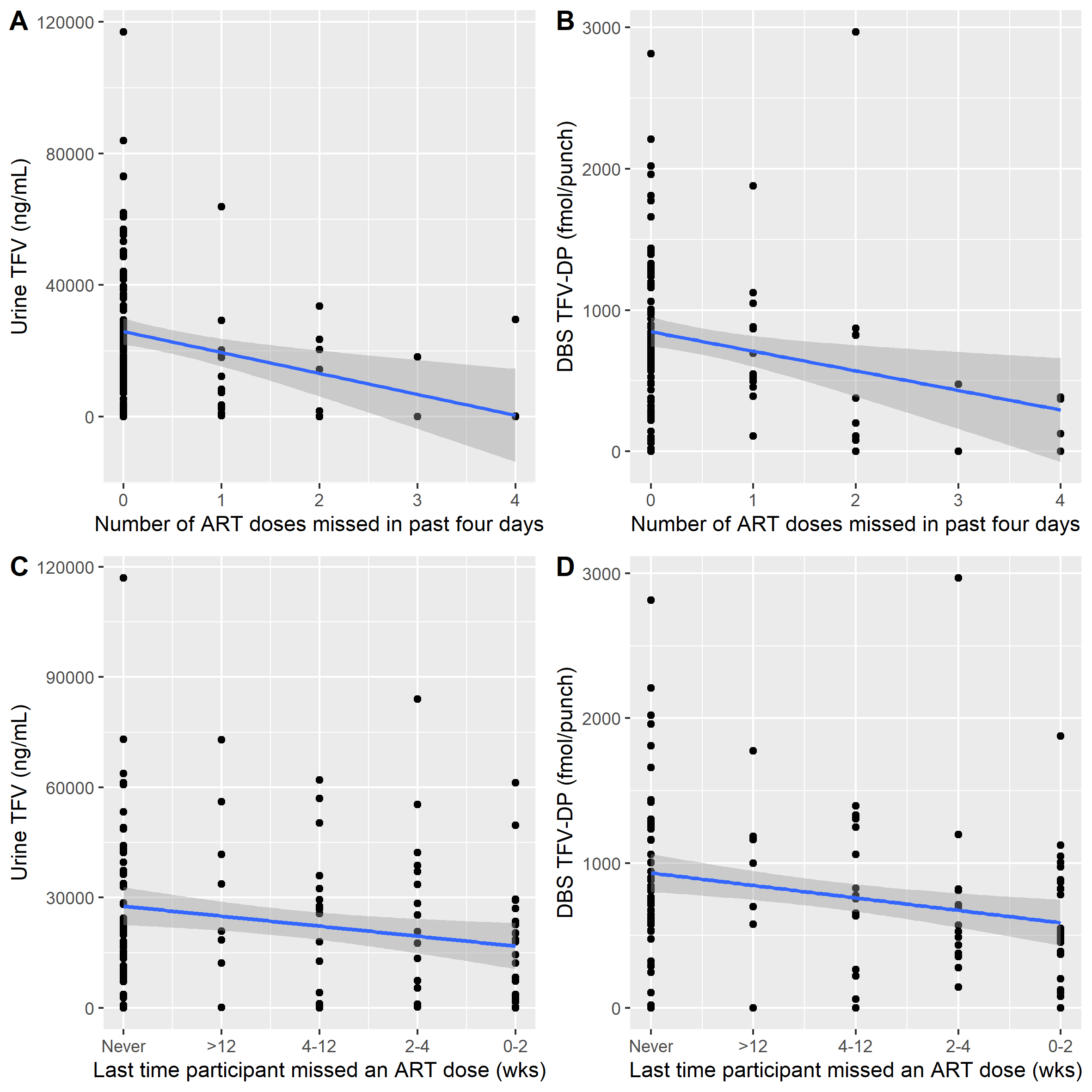

### 6. Table S1 Baseline characteristics of study population, n= 124

| **Variable** | **Levels** | **Total** |
| --- | --- | --- |
| Age, years | Median (IQR) | 39.0 (34.0 to 45.0) |
| Gender | Female | 68 (54.8) |
|  | Male | 56 (45.2) |
| Ethnicity | Black African | 121 (97.6) |
|  | Coloured | 1 (0.8) |
|  | Other | 2 (1.6) |
| Time since ART initiation, years | Median (IQR) | 4.1 (1.5 to 6.1) |
| Current ART regimen | TDF / FTC / EFV | 74 (59.7) |
|  | TDF / 3TC / DTG | 50 (40.3) |
| Time on current regimen, years | Median (IQR) | 1.6 (0.7 to 5.0) |
| ART side effects | No | 120 (96.8) |
|  | Yes | 4 (3.2) |
| Enrolment CD4 count category, cells/µL | <200 | 21 (16.9) |
|  | 200-349 | 25 (20.2) |
|  | 350-499 | 29 (23.4) |
|  | >=500 | 49 (39.5) |
| Last time participant missed a dose of ART | <2 weeks | 30 (24.2) |
|  | 2-4 weeks | 16 (12.9) |
|  | 1-3 months | 16 (12.9) |
|  | >3 months | 8 (6.5) |
|  | Never | 54 (43.5) |
| Number of ART doses missed in past 4 days | 0 | 95 (76.6) |
|  | 1 | 14 (11.3) |
|  | 2 | 9 (7.3) |
|  | 3 | 2 (1.6) |
|  | 4 | 4 (3.2) |
| Urine tenofovir concentration, ng/mL | Median (IQR) | 20000 (7280 to 33625) |
| Dried blood spot tenofovir diphosphate concentration, fmol/punch | Median (IQR) | 734 (471 to 1015) |
| Enrolment viral load, copies/mL | <50 | 57 (46.0) |
|  | 50-999 | 23 (18.5) |
|  | ≥1000 | 44 (35.5) |
| Any HIV drug resistance against current regimen? | No | 19 (15.3) |
|  | Yes | 24 (19.4) |
|  | Unsuccessful | 1 (0.8) |
|  | Viral load <1000 copies/mL | 80 (64.5) |

### 7. Table S2: Sensitivity and specificity of different urine tenofovir and dried blood spot tenofovir diphosphate concentrations to detect viraemia

| **DTG** | | | | | | | | | | | | |
| --- | --- | --- | --- | --- | --- | --- | --- | --- | --- | --- | --- | --- |
| **1000 copies/mL** | | | | | | **50 copies/mL** | | | | | | |
| **Urine TFV** | | | **DBS TFV-DP** | | | **Urine TFV** | | | | **DBS TFV-DP** | | |
| **Urine TFV, ng/mL** | **Sens (%)** | **Spec (%)** | **Urine TFV, ng/mL** | **Sens (%)** | **Spec (%)** | **DBS TFV-DP, fmol/**  **punch** | **Sens (%)** | **Spec (%)** | **DBS TFV-DP, fmol/**  **punch** | | **Sens (%)** | **Spec (%)** |
| Inf | 100.0 | 0.0 | Inf | 100.0 | 0.0 | Inf | 100.0 | 0.0 | Inf | | 100.0 | 0.0 |
| 95050 | 100.0 | 3.0 | 1770 | 100.0 | 3.0 | 95050 | 100.0 | 5.9 | 1770 | | 97.0 | 0.0 |
| 73000 | 100.0 | 6.1 | 1290 | 100.0 | 6.1 | 73000 | 97.0 | 5.9 | 1290 | | 97.0 | 5.9 |
| 67050 | 100.0 | 9.1 | 1224 | 100.0 | 9.1 | 67050 | 97.0 | 11.8 | 1224 | | 97.0 | 11.8 |
| 59100 | 94.1 | 9.1 | 1179 | 100.0 | 12.1 | 59100 | 93.9 | 11.8 | 1179 | | 97.0 | 17.6 |
| 56150 | 94.1 | 12.1 | 1109 | 100.0 | 15.2 | 56150 | 93.9 | 17.6 | 1109 | | 97.0 | 23.5 |
| 52800 | 94.1 | 15.2 | 1029 | 100.0 | 18.2 | 52800 | 90.9 | 17.6 | 1029 | | 97.0 | 29.4 |
| 49650 | 94.1 | 18.2 | 969 | 100.0 | 21.2 | 49650 | 87.9 | 17.6 | 969 | | 97.0 | 35.3 |
| 46050 | 94.1 | 21.2 | 916 | 100.0 | 24.2 | 46050 | 84.8 | 17.6 | 916 | | 97.0 | 41.2 |
| 42700 | 88.2 | 21.2 | 890 | 94.1 | 24.2 | 42700 | 81.8 | 17.6 | 890 | | 93.9 | 41.2 |
| 42050 | 88.2 | 24.2 | 879 | 94.1 | 27.3 | 42050 | 78.8 | 17.6 | 879 | | 93.9 | 47.1 |
| 40700 | 88.2 | 27.3 | 845 | 94.1 | 30.3 | 40700 | 78.8 | 23.5 | 845 | | 93.9 | 52.9 |
| 39150 | 88.2 | 30.3 | 818 | 94.1 | 33.3 | 39150 | 78.8 | 29.4 | 818 | | 90.9 | 52.9 |
| 36250 | 88.2 | 33.3 | 813 | 94.1 | 36.4 | 36250 | 78.8 | 35.3 | 813 | | 87.9 | 52.9 |
| 33750 | 82.4 | 36.4 | 810 | 94.1 | 39.4 | 33750 | 75.8 | 41.2 | 810 | | 87.9 | 58.8 |
| 33350 | 82.4 | 39.4 | 795 | 94.1 | 42.4 | 33350 | 72.7 | 41.2 | 795 | | 87.9 | 64.7 |
| 30700 | 82.4 | 42.4 | 768 | 94.1 | 45.5 | 30700 | 69.7 | 41.2 | 768 | | 84.8 | 64.7 |
| 27050 | 82.4 | 45.5 | 742 | 94.1 | 48.5 | 27050 | 66.7 | 41.2 | 742 | | 81.8 | 64.7 |
| 24400 | 82.4 | 48.5 | 715 | 94.1 | 51.5 | 24400 | 63.6 | 41.2 | 715 | | 81.8 | 70.6 |
| 22850 | 82.4 | 51.5 | 686 | 94.1 | 54.5 | 22850 | 63.6 | 47.1 | **686** | | **81.8** | **76.5** |
| 21650 | 82.4 | 54.5 | 663 | 94.1 | 57.6 | 21650 | 60.6 | 47.1 | 663 | | 78.8 | 76.5 |
| 20650 | 82.4 | 57.6 | 649 | 94.1 | 60.6 | 20650 | 60.6 | 52.9 | 649 | | 75.8 | 76.5 |
| 20400 | 82.4 | 60.6 | 617 | 94.1 | 63.6 | 20400 | 60.6 | 58.8 | 617 | | 75.8 | 82.4 |
| 19400 | 82.4 | 63.6 | 583 | 94.1 | 66.7 | 19400 | 57.6 | 58.8 | 583 | | 72.7 | 82.4 |
| 18100 | 82.4 | 66.7 | 575 | 94.1 | 69.7 | 18100 | 57.6 | 64.7 | 575 | | 69.7 | 82.4 |
| 17300 | 82.4 | 69.7 | 560 | 88.2 | 69.7 | 17300 | 54.5 | 64.7 | 560 | | 66.7 | 82.4 |
| 16000 | 82.4 | 72.7 | 536 | 88.2 | 72.7 | **16000** | **54.5** | **70.6** | 536 | | 66.7 | 88.2 |
| 14250 | 76.5 | 72.7 | 523 | 88.2 | 75.8 | 14250 | 51.5 | 70.6 | 523 | | 63.6 | 88.2 |
| 12450 | 76.5 | 78.8 | 509 | 88.2 | 78.8 | 12450 | 48.5 | 76.5 | 509 | | 60.6 | 88.2 |
| 10950 | 76.5 | 81.8 | 493 | 88.2 | 81.8 | 10950 | 48.5 | 82.4 | 493 | | 57.6 | 88.2 |
| 9740 | 76.5 | 84.8 | **483** | **88.2** | **84.8** | 9740 | 45.5 | 82.4 | 483 | | 54.5 | 88.2 |
| 8160 | 76.5 | 87.9 | 465 | 82.4 | 84.8 | 8160 | 45.5 | 88.2 | 465 | | 51.5 | 88.2 |
| 5495 | 76.5 | 90.9 | 415 | 76.5 | 84.8 | 5495 | 45.5 | 94.1 | 415 | | 48.5 | 88.2 |
| **3495** | **76.5** | **93.9** | 374 | 70.6 | 87.9 | 3495 | 45.5 | 100.0 | 374 | | 45.5 | 94.1 |
| 3065 | 70.6 | 93.9 | 362 | 64.7 | 87.9 | 3065 | 42.4 | 100.0 | 362 | | 42.4 | 94.1 |
| 2610 | 64.7 | 93.9 | 338 | 64.7 | 90.9 | 2610 | 39.4 | 100.0 | 338 | | 39.4 | 94.1 |
| 1521.5 | 58.8 | 93.9 | 304 | 64.7 | 93.9 | 1521.5 | 36.4 | 100.0 | 304 | | 39.4 | 100.0 |
| 432.5 | 52.9 | 93.9 | 266 | 58.8 | 93.9 | 432.5 | 33.3 | 100.0 | 266 | | 36.4 | 100.0 |
| 244 | 52.9 | 97.0 | 194 | 58.8 | 97.0 | 244 | 30.3 | 100.0 | 194 | | 33.3 | 100.0 |
| 221.5 | 52.9 | 100.0 | 125 | 58.8 | 100.0 | 221.5 | 27.3 | 100.0 | 125 | | 30.3 | 100.0 |
| 188 | 47.1 | 100.0 | 106 | 52.9 | 100.0 | 188 | 24.2 | 100.0 | 106 | | 27.3 | 100.0 |
| 160.5 | 41.2 | 100.0 | 92 | 47.1 | 100.0 | 160.5 | 21.2 | 100.0 | 92 | | 24.2 | 100.0 |
| 76 | 35.3 | 100.0 | 80 | 41.2 | 100.0 | 76 | 18.2 | 100.0 | 80 | | 21.2 | 100.0 |
| -Inf | 0.0 | 100.0 | 70 | 35.3 | 100.0 | -Inf | 0.0 | 100.0 | 70 | | 18.2 | 100.0 |
|  |  |  | 40 | 29.4 | 100.0 |  |  |  | 40 | | 15.2 | 100.0 |
|  |  |  | 10 | 23.5 | 100.0 |  |  |  | 10 | | 12.1 | 100.0 |
|  |  |  | -Inf | 0.0 | 100.0 |  |  |  | -Inf | | 0.0 | 100.0 |
| **EFV** | | | | | | | | | | | | |
| **1000 copies/mL** | | | | | | **50 copies/mL** | | | | | | |
| **Urine TFV** | | | **DBS TFV-DP** | | | **Urine TFV** | | | **DBS TFV-DP** | | | |
| **Urine TFV, ng/mL** | **Sens (%)** | **Spec (%)** | **Urine TFV, ng/mL** | **Sens (%)** | **Spec (%)** | **DBS TFV-DP, fmol/**  **punch** | **Sens (%)** | **Spec (%)** | **DBS TFV-DP, fmol/**  **punch** | | **Sens** | **Spec** |
| Inf | 100.0 | 0.0 | Inf | 100.0 | 0.0 | Inf | 100.0 | 0.0 | Inf | | 100.0 | 0.0 |
| 73900 | 100.0 | 2.1 | 2892 | 96.3 | 0.0 | 73900 | 100.0 | 2.5 | 2892 | | 97.1 | 0.0 |
| 62900 | 96.3 | 2.1 | 2417 | 96.3 | 2.1 | 62900 | 97.1 | 2.5 | 2417 | | 97.1 | 2.5 |
| 61650 | 92.6 | 2.1 | 1990 | 96.3 | 4.3 | 61650 | 94.1 | 2.5 | 1990 | | 94.1 | 2.5 |
| 61050 | 92.6 | 4.3 | 1919 | 96.3 | 6.4 | 61050 | 94.1 | 5.0 | 1919 | | 94.1 | 5.0 |
| 58450 | 92.6 | 6.4 | 1844 | 96.3 | 8.5 | 58450 | 94.1 | 7.5 | 1844 | | 94.1 | 7.5 |
| 54700 | 92.6 | 8.5 | 1792 | 96.3 | 10.6 | 54700 | 94.1 | 10.0 | 1792 | | 94.1 | 10.0 |
| 51500 | 92.6 | 10.6 | 1716 | 96.3 | 12.8 | 51500 | 91.2 | 10.0 | 1716 | | 94.1 | 12.5 |
| 49200 | 92.6 | 12.8 | 1547 | 96.3 | 14.9 | 49200 | 91.2 | 12.5 | 1547 | | 94.1 | 15.0 |
| 46400 | 92.6 | 14.9 | 1433 | 92.6 | 14.9 | 46400 | 91.2 | 15.0 | 1433 | | 91.2 | 15.0 |
| 43200 | 88.9 | 14.9 | 1425 | 92.6 | 17.0 | 43200 | 88.2 | 15.0 | 1425 | | 91.2 | 17.5 |
| 39800 | 85.2 | 14.9 | 1407 | 88.9 | 17.0 | 39800 | 85.3 | 15.0 | 1407 | | 88.2 | 17.5 |
| 37200 | 85.2 | 17.0 | 1349 | 88.9 | 19.1 | 37200 | 82.4 | 15.0 | 1349 | | 88.2 | 20.0 |
| 36700 | 85.2 | 19.1 | 1303 | 88.9 | 21.3 | 36700 | 82.4 | 17.5 | 1303 | | 88.2 | 22.5 |
| 36150 | 85.2 | 21.3 | 1293 | 88.9 | 23.4 | 36150 | 82.4 | 20.0 | 1293 | | 88.2 | 25.0 |
| 34800 | 85.2 | 23.4 | 1277 | 88.9 | 25.5 | 34800 | 82.4 | 22.5 | 1277 | | 85.3 | 25.0 |
| 33550 | 81.5 | 23.4 | 1257 | 88.9 | 27.7 | 33550 | 79.4 | 22.5 | 1257 | | 85.3 | 27.5 |
| 32950 | 77.8 | 23.4 | 1241 | 88.9 | 29.8 | 32950 | 76.5 | 22.5 | 1241 | | 85.3 | 30.0 |
| 30950 | 77.8 | 25.5 | 1211 | 88.9 | 31.9 | 30950 | 76.5 | 25.0 | 1211 | | 85.3 | 32.5 |
| 29450 | 74.1 | 25.5 | 1183 | 88.9 | 34.0 | 29450 | 73.5 | 25.0 | 1183 | | 85.3 | 35.0 |
| 29350 | 74.1 | 27.7 | 1172 | 88.9 | 36.2 | 29350 | 73.5 | 27.5 | 1172 | | 82.4 | 35.0 |
| 28900 | 70.4 | 27.7 | 1143 | 88.9 | 40.4 | 28900 | 70.6 | 27.5 | 1143 | | 79.4 | 37.5 |
| 28350 | 70.4 | 29.8 | 1092 | 88.9 | 42.6 | 28350 | 70.6 | 30.0 | 1092 | | 79.4 | 40.0 |
| 27950 | 66.7 | 29.8 | 1053 | 88.9 | 44.7 | 27950 | 67.6 | 30.0 | 1053 | | 79.4 | 42.5 |
| 27350 | 66.7 | 31.9 | 1026 | 85.2 | 44.7 | 27350 | 67.6 | 32.5 | 1026 | | 76.5 | 42.5 |
| 26950 | 66.7 | 34.0 | 1003 | 81.5 | 46.8 | 26950 | 67.6 | 35.0 | 1003 | | 73.5 | 45.0 |
| 26100 | 63.0 | 34.0 | 987 | 81.5 | 48.9 | 26100 | 64.7 | 35.0 | 987 | | 73.5 | 47.5 |
| 24850 | 63.0 | 36.2 | 935 | 81.5 | 51.1 | 24850 | 64.7 | 37.5 | 935 | | 73.5 | 50.0 |
| 24150 | 63.0 | 38.3 | 897 | 81.5 | 53.2 | 24150 | 64.7 | 40.0 | 897 | | 73.5 | 52.5 |
| 23700 | 63.0 | 40.4 | 889 | 81.5 | 55.3 | 23700 | 64.7 | 42.5 | 889 | | 73.5 | 55.0 |
| 23350 | 63.0 | 42.6 | 881 | 81.5 | 57.4 | 23350 | 64.7 | 45.0 | 881 | | 73.5 | 57.5 |
| 22950 | 63.0 | 44.7 | 876 | 81.5 | 59.6 | 22950 | 64.7 | 47.5 | 876 | | 70.6 | 57.5 |
| 22550 | 59.3 | 44.7 | 858 | 81.5 | 61.7 | 22550 | 61.8 | 47.5 | 858 | | 70.6 | 60.0 |
| 22150 | 59.3 | 46.8 | 839 | 77.8 | 63.8 | 22150 | 61.8 | 50.0 | 839 | | 67.6 | 62.5 |
| 21750 | 59.3 | 48.9 | 831 | 77.8 | 66.0 | 21750 | 61.8 | 52.5 | 831 | | 67.6 | 65.0 |
| 21300 | 59.3 | 51.1 | 826 | 74.1 | 66.0 | 21300 | 58.8 | 52.5 | 826 | | 64.7 | 65.0 |
| 20950 | 59.3 | 53.2 | 823 | 74.1 | 68.1 | 20950 | 58.8 | 55.0 | 823 | | 64.7 | 67.5 |
| 20650 | 59.3 | 55.3 | **816** | **74.1** | **70.2** | 20650 | 58.8 | 57.5 | **816** | | **64.7** | **70.0** |
| **20100** | **59.3** | **57.4** | 794 | 70.4 | 70.2 | **20100** | **58.8** | **60.0** | 794 | | 61.8 | 70.0 |
| 19150 | 55.6 | 57.4 | 771 | 66.7 | 70.2 | 19150 | 55.9 | 60.0 | 771 | | 58.8 | 70.0 |
| 18350 | 55.6 | 59.6 | 758 | 63.0 | 70.2 | 18350 | 55.9 | 62.5 | 758 | | 55.9 | 70.0 |
| 18150 | 51.9 | 59.6 | 744 | 63.0 | 72.3 | 18150 | 52.9 | 62.5 | 744 | | 52.9 | 70.0 |
| 18050 | 48.1 | 59.6 | 726 | 63.0 | 74.5 | 18050 | 50.0 | 62.5 | 726 | | 52.9 | 72.5 |
| 17800 | 48.1 | 63.8 | 713 | 59.3 | 74.5 | 17800 | 50.0 | 67.5 | 713 | | 50.0 | 72.5 |
| 17050 | 48.1 | 66.0 | 704 | 55.6 | 76.6 | 17050 | 47.1 | 67.5 | 704 | | 47.1 | 75.0 |
| 15900 | 44.4 | 66.0 | 694 | 51.9 | 76.6 | 15900 | 44.1 | 67.5 | 694 | | 44.1 | 75.0 |
| 14850 | 44.4 | 68.1 | 684 | 51.9 | 78.7 | 14850 | 44.1 | 70.0 | 684 | | 44.1 | 77.5 |
| 14200 | 40.7 | 68.1 | 663 | 51.9 | 80.9 | 14200 | 41.2 | 70.0 | 663 | | 44.1 | 80.0 |
| 13350 | 40.7 | 70.2 | 647 | 51.9 | 83.0 | 13350 | 41.2 | 72.5 | 647 | | 44.1 | 82.5 |
| 12450 | 40.7 | 72.3 | 639 | 51.9 | 85.1 | 12450 | 41.2 | 75.0 | 639 | | 44.1 | 85.0 |
| 11400 | 40.7 | 76.6 | 626 | 48.1 | 85.1 | 11400 | 38.2 | 77.5 | 626 | | 41.2 | 85.0 |
| 10200 | 40.7 | 78.7 | 607 | 48.1 | 87.2 | 10200 | 38.2 | 80.0 | 607 | | 41.2 | 87.5 |
| 9105 | 40.7 | 80.9 | 583 | 44.4 | 87.2 | 9105 | 38.2 | 82.5 | 583 | | 38.2 | 87.5 |
| 8340 | 40.7 | 83.0 | 552 | 44.4 | 89.4 | 8340 | 38.2 | 85.0 | 552 | | 38.2 | 90.0 |
| 8165 | 37.0 | 83.0 | 531 | 40.7 | 89.4 | 8165 | 35.3 | 85.0 | 531 | | 35.3 | 90.0 |
| 8020 | 37.0 | 85.1 | 514 | 37.0 | 89.4 | 8020 | 35.3 | 87.5 | 514 | | 32.4 | 90.0 |
| 7665 | 37.0 | 87.2 | 491 | 37.0 | 91.5 | 7665 | 35.3 | 90.0 | 491 | | 32.4 | 92.5 |
| 7225 | 33.3 | 87.2 | 480 | 37.0 | 93.6 | 7225 | 32.4 | 90.0 | 480 | | 29.4 | 92.5 |
| 6255 | 33.3 | 89.4 | 456 | 33.3 | 93.6 | 6255 | 29.4 | 90.0 | 456 | | 26.5 | 92.5 |
| 4745 | 33.3 | 91.5 | 412 | 33.3 | 95.7 | 4745 | 29.4 | 92.5 | 412 | | 26.5 | 95.0 |
| 3825 | 29.6 | 91.5 | 386 | 29.6 | 95.7 | 3825 | 26.5 | 92.5 | 386 | | 23.5 | 95.0 |
| 2830 | 29.6 | 93.6 | 338 | 25.9 | 95.7 | 2830 | 26.5 | 95.0 | 338 | | 20.6 | 95.0 |
| 1855 | 29.6 | 95.7 | 285 | 25.9 | 97.9 | 1855 | 23.5 | 95.0 | 285 | | 20.6 | 97.5 |
| 1385 | 29.6 | 97.9 | 270 | 22.2 | 97.9 | 1385 | 23.5 | 97.5 | 270 | | 17.6 | 97.5 |
| 1090 | 25.9 | 97.9 | 241 | 18.5 | 97.9 | 1090 | 20.6 | 97.5 | 241 | | 14.7 | 97.5 |
| 987 | 22.2 | 97.9 | 210 | 14.8 | 97.9 | 987 | 17.6 | 97.5 | 210 | | 11.8 | 97.5 |
| 820.5 | 18.5 | 97.9 | 163 | 14.8 | 100.0 | 820.5 | 14.7 | 97.5 | 163 | | 11.8 | 100.0 |
| 692 | 14.8 | 97.9 | 117 | 11.1 | 100.0 | 692 | 11.8 | 97.5 | 117 | | 8.8 | 100.0 |
| 515 | 11.1 | 97.9 | 55 | 7.4 | 100.0 | 515 | 8.8 | 97.5 | 55 | | 5.9 | 100.0 |
| 224 | 7.4 | 97.9 | -Inf | 0.0 | 100.0 | 224 | 5.9 | 97.5 | -Inf | | 0.0 | 100.0 |
| 52.5 | 3.7 | 97.9 |  |  |  | 52.5 | 2.9 | 97.5 |  | |  |  |
| -Inf | 0.0 | 100.0 |  |  |  | -Inf | 0.0 | 100.0 |  | |  |  |

### 8. Table S3: Median urine TFV and DBS TFV-DP by presence of HIV drug resistance

|  | **No HIV drug resistance** | **HIV drug resistance** |
| --- | --- | --- |
| All (n = 43)* | N = 19 | N = 24 |
| Urine TFV ng/mL, Median (IQR) | 343 (0-20950) | 17300 (1120-29350) |
| DBS TFV-DP fmol/punch, Median (IQR) | 103 (10-374) | 646 (388-820) |
| EFV (n = 28) | N = 4 | N = 24 |
| Urine TFV ng/mL, Median (IQR) | 15490 (3146-35675) | 18100 (1090-29400) |
| DBS TFV-DP fmol/punch, Median (IQR) | 370 (219-634) | 696 (386-828) |
| DTG (n = 14) | N = 15 | N = 1 |
| Urine TFV ng/mL, Median (IQR) | 169 (0-8715) | 3340 (3340-3340) |
| DBS TFV-DP fmol/punch, Median (IQR) | 80 (10-328) | 454 (454-454) |

*Numbers low for meaningful hypothesis testing

### 9. Table S4: Self-reported adherence compared to urine tenofovir and dried blood spot tenofovir diphosphate concentrations

| **Adherence variable** | | **Urine TFV concentration** | | | **DBS TFV-DP** | |
| --- | --- | --- | --- | --- | --- | --- |
|  |  | **Median (IQR) (ng/mL)** | **Coefficient^†^ (95% CI), P, r^2^** | **Median (IQR) (fmol/punch)** | | **Coefficient^†^ (95% CI) , P, r^2^** |
| Number of ART doses missed in past 4 days | 0 | 22600 (11000-37200) | -6380 (-10221 to -2538)  P = 0.001  r^2^ = 0.08 | 783 (572-1161) | | -139 (-238 to -39)  P = 0.007  r^2^ = 0.06 |
|  | 1 | 8125 (3398-18450) |  | 541 (497-879) | |  |
|  | 2 | 1620 (0-20400) |  | 377 (109-824) | |  |
|  | 3 | 9100 (4550-13650) |  | 238 (119-357) | |  |
|  | 4 | 128 (79-7489) |  | 247 (93-374) | |  |
| Last time participant missed a dose of ART (weeks) | Never | 21750 (10800-35675) | -2724 (-4911 to -537)  P = 0.015  r^2^ = 0.05 | 839 (601-1218) | | -86 (-141 to -31)  P = 0.003  r^2^ = 0.07 |
|  | >12 | 27300 (16925-45375) |  | 1080 (668-1183) | |  |
|  | 4-12 | 26300 (3348-33300) |  | 704 (542-1106) | |  |
|  | 2-4 | 23000 (6865-37500) |  | 631 (420-812) | |  |
|  | <2 | 8125 (582-20350) |  | 489 (244-858) | |  |

**^†^**Linear regression models
